# Epidemiological parameters of coronavirus disease 2019: a pooled analysis of publicly reported individual data of 1155 cases from seven countries

**DOI:** 10.1101/2020.03.21.20040329

**Authors:** Shujuan Ma, Jiayue Zhang, Minyan Zeng, Qingping Yun, Wei Guo, Yixiang Zheng, Shi Zhao, Maggie H. Wang, Zuyao Yang

## Abstract

**Background:** The outbreak of coronavirus disease 2019 (COVID-19) has been declared a pandemic by the World Health Organization, while several key epidemiological parameters of the disease remain to be clarified. This study aimed to obtain robust estimates of the incubation period, upper limit of latent period (interval between infector’s exposure and infectee’s exposure), serial interval, time point of exposure (the day of infectee’s exposure to infector relative to the latter’s symptom onset date) and basic reproduction number (R_0_) of COVID-19.

**Methods:** Between late February and early March of 2020, the individual data of laboratory confirmed cases of COVID-19 were retrieved from 10728 publicly available reports released by the health authorities of and outside China and from 1790 publications identified in PubMed and CNKI. To be eligible, a report had to contain the data that allowed for estimation of at least one parameter. As relevant data mainly came from clustering cases, the clusters for which no evidence was available to establish transmission order were all excluded to ensure accuracy of estimates. Additionally, only the cases with an exposure period spanning 3 days or less were included in the estimation of parameters involving exposure date, and a simple method for determining exposure date was adopted to ensure the error of estimates be small (< 0.3 day). Depending on specific parameters, three or four of normal, lognormal, Weibull, and gamma distributions were fitted to the datasets and the results from appropriate models were presented.

**Findings:** In total, 1155 cases from China, Japan, Singapore, South Korea, Vietnam, Germany and Malaysia were included for the final analysis. The mean and standard deviation were 7.44 days and 4.39 days for incubation period, 2.52 days and 3.95 days for the upper limit of latent period, 6.70 days and 5.20 days for serial interval, and −0.19 day (i.e., 0.19 day before infector’s symptom onset) and 3.32 days for time point of exposure. R_0_ was estimated to be 1.70 and 1.78 based on two different formulas. For 39 (6.64%) cases, the incubation periods were longer than 14 days. In 102 (43.78%) infector-infectee pairs, transmission occurred before infectors’ symptom onsets. In 27 (3.92%) infector-infectee pairs, infectees’ symptom onsets occurred before those of infectors. Stratified analysis showed that incubation period and serial interval were consistently longer for those with less severe disease and for those whose primary cases had less severe disease. Asymptomatic transmission was also observed.

**Interpretation:** This study obtained robust estimates of several key epidemiological parameters of COVID-19. The findings support current practice of 14-day quarantine of persons with potential exposure, but also suggest that longer monitoring periods might be needed for selected groups. The estimates of serial interval, time point of exposure and latent period provide consistent evidence on pre-symptomatic transmission. This together with asymptomatic transmission and the generally longer incubation and serial interval of less severe cases suggests a high risk of long-term epidemic in the absence of appropriate control measures.

**Funding:** This work received no funding from any source.

## Introduction

In December 2019, a novel enveloped RNA beta-coronavirus, which was later named severe acute respiratory syndrome coronavirus 2 (SARS-CoV-2), emerged in Wuhan, the capital city of Hubei province of China.^1^ The disease caused by SARS-CoV-2, i.e., coronavirus disease 2019 (COVID-19), spread across and outside China rapidly, and was declared a pandemic by the World Health Organization on 11 March 2020.

Despite the explosive growth of the number of studies on COVID-19, several key epidemiological parameters of the disease remain to be clarified, among which are incubation period and serial interval. The two parameters have important implications for the monitoring, surveillance and control of infectious disease. The mean or median incubation period and serial interval estimated by previous studies were mostly 4 to 5 days,^2-7^ and 4 to 4.5 days,^8-12^ respectively. While some of the studies included only a limited number of cases (around or less than 100),^2,4,6,7,9-12^ others might have suffered from inaccuracy of original data. For example, in some studies, most cases had a long or even unclear interval of exposure, making it difficult to determine the exact exposure date^3,5,13^ and giving rise to error. Besides, the order of transmission (i.e., who is infector and who is infectee) in clustering cases, which is crucial to estimation of both parameters, is easy to be mistaken given the possibility of pre-symptomatic and asymptomatic transmission,^14-17^ but previous studies rarely described how this issue was handled.

Incubation period and serial interval are also key parameters for epidemiological modelling in predicting the transmission dynamics, including the basic reproduction number (R_0_). R_0_ is defined as the average number of secondary cases caused by a single infectious individual in a totally susceptible population.^18^ Estimation of R_0_ may require the knowledge of latent period and infectious period,^19,20^ which are both difficult to measure directly. For a disease that is not infectious until onset of symptoms, latent period is equivalent to incubation period, and infectious period can commonly be approximated by the difference of serial interval minus incubation period.^18^ However, for COVID-19, the latent period appears to be shorter than incubation period as pre-symptomatic transmission might occur,^16,17^ and this should be accounted for when estimating R_0_. To our best knowledge, no published studies estimated the latent period of COVID-19, or how often and approximately at which time point the disease could be transmitted prior to the symptomatic onset.

This study made use of the large amount of data reported by the health authorities of and outside China and those from published studies to address the above issues. Specifically, it was aimed to obtain robust estimates of the following epidemiological parameters of COVID-19: 1) incubation period, 2) the upper limit of latent period, as the exact latent period cannot be observed, 3) serial interval, 4) time point of exposure, referring to the day of infectee’s exposure to infector relative to the latter’s symptom onset date, and 5) R_0_.

## Methods

### Data sources

For China, all provinces, autonomous regions and municipalities (including mainland, Hong Kong, Macau and Taiwan) that had reported cases of COVID-19 were identified according to the daily updates by the National Health Commission of China.^21^ Then, the official websites and WeChat accounts (if any) of local governments and health authorities (e.g., Municipal Health Commission, Center for Disease Control and Prevention, Department of Health) were checked manually through February 20, 2020 for mainland China and March 3, 2020 for Hong Kong, Macau and Taiwan to identify and download the reports on laboratory-confirmed cases of COVID-19. Google and Baidu were searched to identify public media reports quoting government departments, which were normally based on official press releases. Chinese words for the following terms were used to do the search: (‘family’ OR ‘household’) AND (‘cluster’ OR ‘dinner’ OR ‘party’) AND ‘infection’.

Other countries reporting COVID-19 cases were identified according to the ‘coronavirus disease (COVID-19) situation reports’ by World Health Organization.^22^ For Japan and Singapore, the information of confirmed cases was retrieved from their respective Ministry of Health. For Malaysia and Vietnam, public media reports quoting government sources were adopted. The dates of search for the four countries varied from February 29 to March 2, 2020. We also searched for individual cases from relevant departments and public media of the United States, the United Kingdom, Canada and Australia, but failed to find any with details allowing for parameters estimation in this study. Typically, the dates of exposure and symptom onset were lacking, e.g., see reference.^23^ Owing to language barrier, we did not do a comprehensive search for other countries. A full list of the abovementioned data sources, including their URLs, can be found in Appendix 1.

PubMed was searched to identify relevant publications by using the following terms: ‘coronavirus’, ‘2019-nCov’, ‘SARS-CoV-2’, and ‘COVID-19’. The search was restricted to December 1 and February 27, 2020. After February 27, we did not update the search but still kept an eye on the newly published studies on COVID-19, which were frequently disseminated by public media and the official WeChat accounts of various academic entities in China, or recommended by experts in this field to the authors of the present study. The China National Knowledge Infrastructure (CNKI) was searched through March 11, 2020 to identify publications in Chinese journals using ‘novel coronavirus’ (‘Xin Xing Guan Zhuang Bing Du’ in Chinese pinyin), which was the official Chinese name for SARS-CoV-2. The reference lists of eligible publications were also checked to see if there were other eligible studies not found by previous searches. A supplementary list of included studies can be found in Appendix 2.

### Definitions and inclusion criteria

To be eligible, a report had to contain individual data that allowed for estimation of at least one of the following parameters of laboratory-confirmed cases of COVID-19: incubation period, serial interval, time point of exposure, and the upper limit of latent period.

Incubation period was defined as the time interval between exposure and onset of disease symptoms. To obtain an accurate estimate, only the cases with an exposure period spanning 3 days or less were included in the analysis. For those exposed for three continuous days and those exposed on two dates with one day apart (i.e., exposed on the first and third days), the second day was uniformly used as the exposure date in estimation. For those exposed for two continuous days, the first day was uniformly used as the exposure date in estimation. This approach ensured the upper limit of error in the estimated incubation period be smaller than 1 day for the cases with a 2-day or 3-day exposure, regardless of when exactly (i.e., first, second, or third day) the transmission actually occurred. In reality, the overall error was bound to be much smaller than 1 day, as most included cases were exposed for only 1 day which would dilute the overall error.

Serial interval was defined as the duration between symptom onset of an infector (e.g., a primary case) and that of an infectee (e.g., a secondary case) in a transmission chain, which was typically seen in cluster infections. A negative value meant that the infectee’s symptoms occurred before the infector’s symptoms. For two cases to qualify as an infector-infectee pair and be included in this study, the following two criteria must both be fulfilled. First, there was evidence that the presumed infector had been exposed outside the cluster (e.g., close contact with a confirmed case, travel history to Hubei, exposure to a person who returned from Hubei) before he/she joined the group activities such as family gathering and business conference that gave rise to cluster infections. Second, the presumed infectee was exposed to the presumed infector only in the 14 days prior to symptom onset, without other exposure histories.

Time point of exposure in this study referred to the day of infectee’s exposure to infector relative to the latter’s symptom onset date, and was estimated by the infectee’s exposure date minus the infector’s symptom onset date, where a negative value meant that the exposure occurred before the infector developed symptoms. The upper limit of latent period was defined as the interval between the exposure of infector and that of infectee in a transmission chain, which was used to estimate the longest possible latent period of an individual infector (the exact latent period cannot be observed in reality). If there were more than one infectee caused by an infector, only the one with the earliest exposure date was used in estimating this parameter. Estimation of the time point of exposure and upper limit of latent period also involved determination of exposure date and judgement about transmission chain, hence the related principles applied in estimating incubation period and serial interval were also followed here.

### Screening, data extraction, and quality control

Six researchers were involved in data collection. The reports retrieved through the above searches were scrutinized one by one according to the inclusion criteria specified above. The following data were extracted from eligible reports by using a standard extraction form which was pilot-tested with the reports from Liaoning province of China: the geographical location concerned, age, sex, type of exposure, first date and period (if applicable) of exposure, date of symptom onset, initial symptoms, severity, whether the case was from a cluster. For a clustering case, the generation he/she belonged to and the exposure date, symptom onset date and severity of the infector were also recorded. The retrieved reports were split into six parts, with each researcher responsible for one part. For each part, the eligibility of and data extracted from reports were firstly determined by one researcher and then cross-checked by another three researchers. All uncertainty and disagreements were discussed among the researchers. If no consensus could be reached after discussion, the concerned cases would be excluded to ensure the correctness and accuracy of data. For example, if it could not be determined who was infected through attending a group activity (i.e., infectee) and who had been infected before he/she attended the activity (i.e., infector), then all clustering cases related to the activity had to be excluded.

### Data analysis

The basic characteristics of included cases were summarized descriptively. The lognormal, Weibull, and gamma distributions were fitted to the datasets of incubation period and upper limit of latent period, and the key parameters were estimated by using the maximum likelihood approach. For serial interval and time point of exposure, we assumed that they followed a normal distribution.^8^ This was to address the negative values in the serial interval reported by previous studies, in which case the lognormal, Weibull, and gamma distributions may be limited.^24,25^ For each parameter, the range, median, selected percentiles, mean and standard deviation were estimated. The 95% confidence intervals (CI) of mean and standard deviation were estimated by using the bootstrap technique. R_0_ was estimated by using two widely accepted formula:^8,9,19,20^

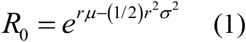

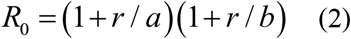

where *r* is exponential growth rate, μ is mean serial interval, σ is standard deviation of serial interval, 1/a is latent period, and 1/b is infectious period. The exponential growth rate was obtained directly from a previous study reporting incidence data of the early stage of epidemic,^1^ while the other parameters came from the present study. The latent period was approximated by the upper limit of latent period, and the infectious period was approximated as serial interval minus the upper limit of latent period. Although the true value of latent period was bound to be smaller than the upper limit, we argued that this would not exert a significant impact on the estimation of R_0_ according to the above formula (2), because when the latent period became shorter the infectious period would become longer accordingly, given a fixed serial interval.

Sensitivity analyses were conducted to examine the robustness of the three parameters involving exposure date. Specifically, the third day for those whose exposure period spanned three days and the second day for those exposed for two continuous days were used as their exposure dates in sensitivity analyses. The difference between the estimates from sensitivity analysis and those from primary analysis represents the largest possible error in the latter. For serial interval and time point of exposure, which had negative values, the datasets were also fitted with shifted lognormal, Weibull and gamma distributions to see if the mean and standard deviation would change much. Based on the new estimates from sensitivity analyses, R_0_ was also re-estimated. All statistical analyses were conducted with SAS software, version 9.4.

## Results

### Process and output of data collection

The process of data collection is summarized in Figure 1. In total, 1155 cases were included for the final analysis, including 1054 (91.3%) from China, 39 from Japan, 37 from Singapore, 11 from South Korea, 7 from Vietnam, 4 from Germany, and 3 from Malaysia. The cases from China covered 151 cities of 30 provinces out of 34 in total. No case from Chongqing, Xinjiang, Tibet and Macau was included in this study.

**Figure 1.**
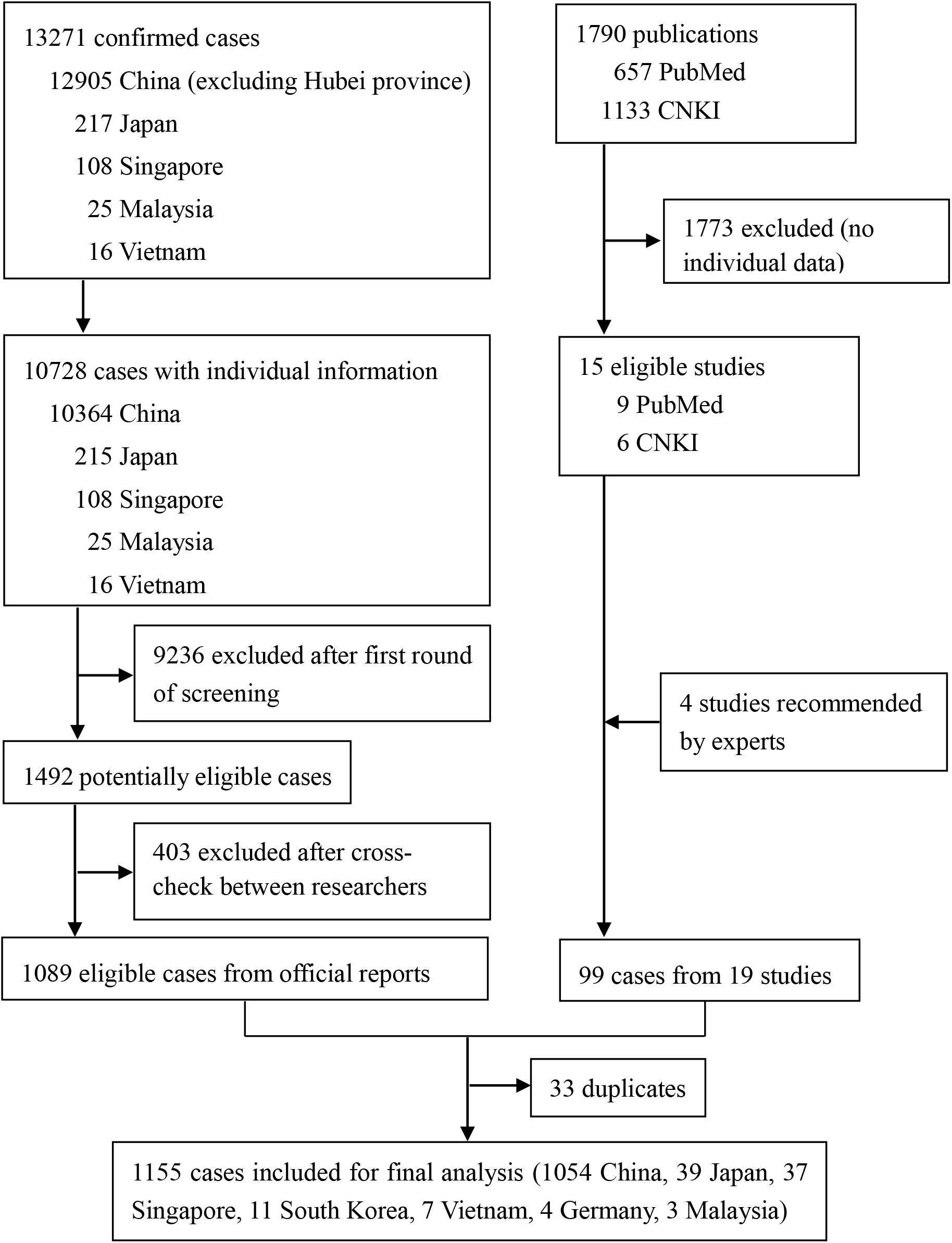
The flowchart of cases selection from the official reports and published studies.

### Characteristics of included cases

The age of included cases ranged from 5 days to 90 years (Table 1). There were 122 cases (12.92%) with a travel history to Hubei province where Wuhan is the capital city. The source of exposure of 887 cases was known confirmed cases. The exposure period was exactly 1 day in 426 (50.3%) cases, spanned 2-3 days in 196 (23.1%) cases, and spanned more than 3 days in the other cases (who were not included in estimation of parameters involving exposure date). Among the cases with relevant information (n=329, 28.48%), 49 (14.89%) were asymptomatic, 256 (77.81%) mild to moderate, and 24 (7.29%) severe. The top five initial symptoms were fever (73.92%), cough (24.06%), fatigue (7.49%), malaise (7.20%), and chills (4.03%). Clustering cases accounted for 81.4% of all, and most of them were the second generation. It should be noted that for each generation there were some cases excluded from this study, because no information was available for estimating the parameters of interest.

**Table 1.** Characteristics of included cases

| Characteristics | No. (%)<sup>*</sup> |
| --- | --- |
| Age (n=1035) |  |
| Range | 5 days, 90 years |
| Mean (standard deviation) (year) | 46.26 (17.19) |
| 0 - 18 | 50 (4.83) |
| 19 - 64 | 833 (80.48) |
| ≥65 | 152 (14.69) |
| Sex (n=1126) |  |
| Male | 547 (48.58) |
| Female | 579 (51.42) |
| Travel history to Hubei (n=944) |  |
| Yes | 122 (12.92) |
| No | 822 (87.08) |
| Infecter was a known confirmed case (n=1155) |  |
| Yes | 887 (76.80) |
| No | 268 (23.20) |
| Span of exposure period (n=847) |  |
| 1 day | 426 (50.30) |
| 2-3 days | 196 (23.14) |
| >3 days | 225 (26.56) |
| Top 5 initial symptoms (n=694) |  |
| Fever | 513 (73.92) |
| Cough | 167 (24.06) |
| Fatigue | 52 (7.49) |
| Malaise | 50 (7.20) |
| Chills | 28 (4.03) |
| Severity (n=329) |  |
| Asymptomatic | 49 (14.89) |
| Mild to moderate | 256 (77.81) |
| Severe | 24 (7.29) |
| Clustering cases (n=1155) |  |
| Yes | 940 (81.39) |
| No | 215 (18.61) |
| Generation of clustering cases (n=929) |  |
| 1<sup>st</sup> | 54 (5.81) |
| 2<sup>nd</sup> | 718 (77.29) |
| 3<sup>rd</sup> | 131 (14.10) |
| 4<sup>th</sup> or higher | 26 (2.8) |

<sup>\*</sup> Unless other specified

### Epidemiological parameters

Incubation period were estimated from 587 cases, of whom 408 were exposed for 1 day and the others for two or three days. The incubation period of individual cases ranged from 0 to 23 days (Table 2), with a median of 7 days and 6.64% (n=39) of them longer than 14 days. The estimates from lognormal, Weibull and gamma models were almost the same. Thus, lognormal model was used uniformly for this parameter (Figure 2(A)). The mean incubation period was estimated at 7.44 days (95% CI: 7.10, 7.78) and the standard deviation 4.39 days (95% CI: 3.97, 4.49).

**Table 2.**
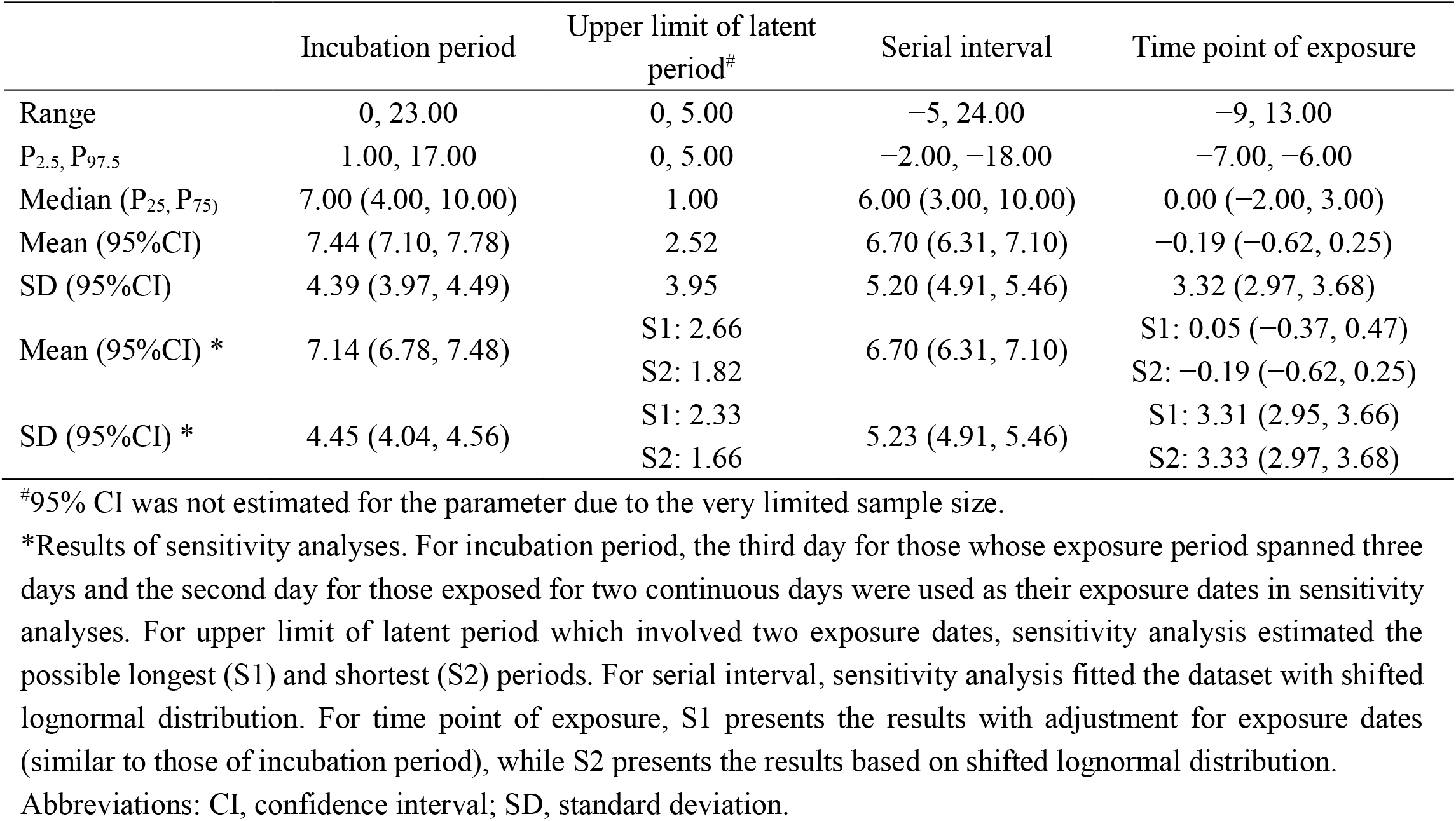
The estimates of incubation period, upper limit of latent period, serial interval and time point of exposure (unit: day)

|  | Incubation period | Upper limit of latent period<sup>#</sup> | Serial interval | Time point of exposure |
| --- | --- | --- | --- | --- |
| Range | 0, 23.00 | 0, 5.00 | −5, 24.00 | −9, 13.00 |
| P<sub>2.5</sub>, P<sub>97.5</sub> | 1.00, 17.00 | 0, 5.00 | −2.00, −18.00 | −7.00, −6.00 |
| Median (P<sub>25</sub>, P<sub>75</sub>) | 7.00 (4.00, 10.00) | 1.00 | 6.00 (3.00, 10.00) | 0.00 (−2.00, 3.00) |
| Mean (95%CI) | 7.44 (7.10, 7.78) | 2.52 | 6.70 (6.31, 7.10) | −0.19 (−0.62, 0.25) |
| SD (95%CI) | 4.39 (3.97, 4.49) | 3.95 | 5.20 (4.91, 5.46) | 3.32 (2.97, 3.68) |
| Mean (95%CI) * | 7.14 (6.78, 7.48) | S1: 2.66 | 6.70 (6.31, 7.10) | S1: 0.05 (−0.37, 0.47) |
|  |  | S2: 1.82 |  | S2: −0.19 (−0.62, 0.25) |
| SD (95%CI) * | 4.45 (4.04, 4.56) | S1: 2.33 | 5.23 (4.91, 5.46) | S1: 3.31 (2.95, 3.66) |
|  |  | S2: 1.66 |  | S2: 3.33 (2.97, 3.68) |

<sup>#</sup>95% CI was not estimated for the parameter due to the very limited sample size.
\*Results of sensitivity analyses. For incubation period, the third day for those whose exposure period spanned three days and the second day for those exposed for two continuous days were used as their exposure dates in sensitivity analyses. For upper limit of latent period which involved two exposure dates, sensitivity analysis estimated the possible longest (S1) and shortest (S2) periods. For serial interval, sensitivity analysis fitted the dataset with shifted lognormal distribution. For time point of exposure, S1 presents the results with adjustment for exposure dates (similar to those of incubation period), while S2 presents the results based on shifted lognormal distribution.
Abbreviations: CI, confidence interval; SD, standard deviation.

**Figure 2.**
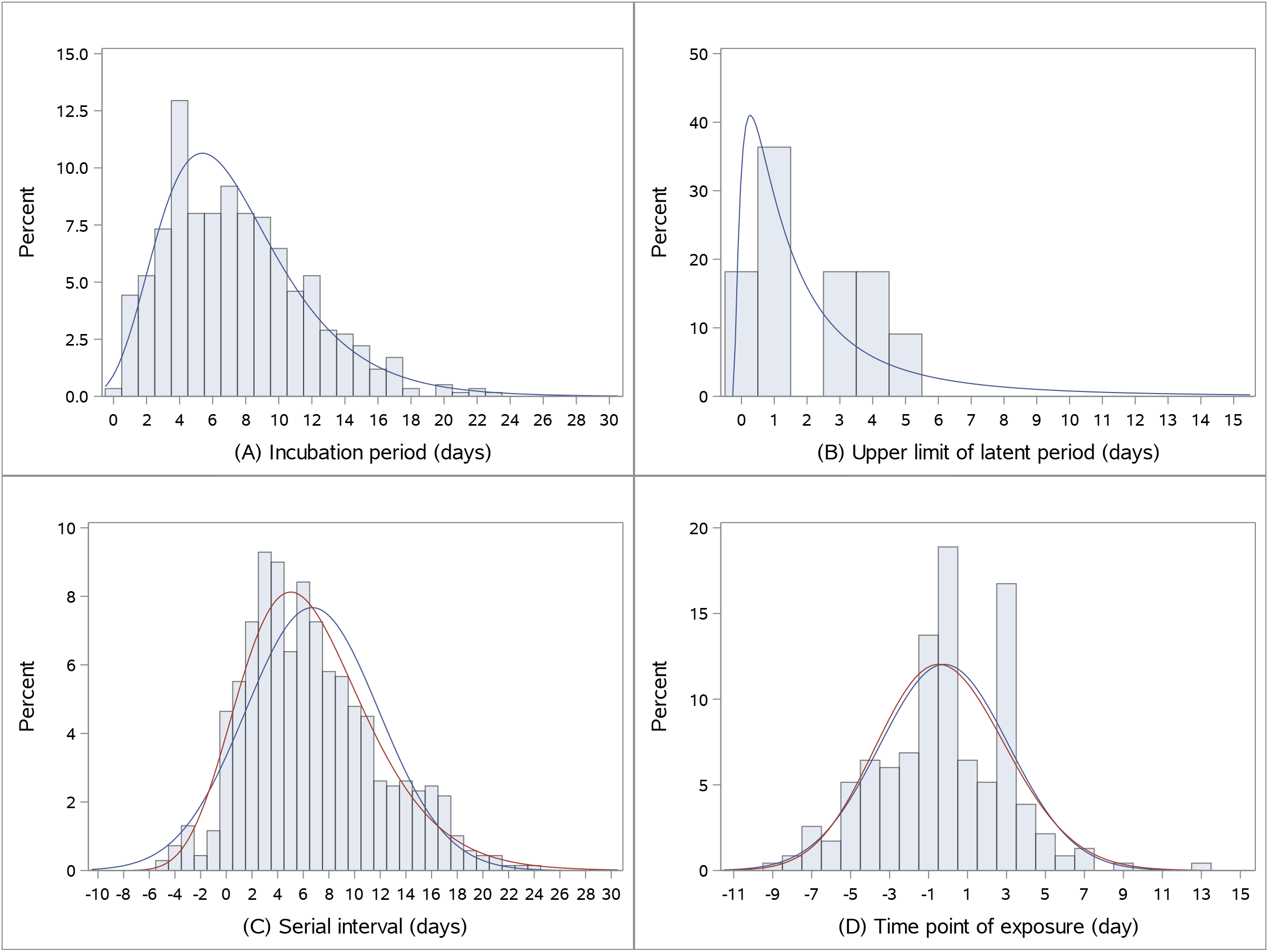
The distribution of (A) incubation period, (B) upper limit of latent period, (C) serial interval, and (D) time point of exposure. The curves of lognormal distribution are presented in all four sub-figures. In (C) and (D), red lines are based on lognormal distribution and blue lines are based normal distribution.

Only 11 pairs of infector-infectee was eligible for estimation of the upper limit of latent period, which ranged from 0 to 5 days (Table 2) with a median of 1 day. The mean and standard deviation derived from lognormal model, which fitted the dataset best (Figure 2(B)), were 2.52 days and 3.95 days, respectively. The small sample size precluded further analysis.

The serial interval was estimated for 689 pairs of infector-infectee and ranged from −5 to 24 days (Table 2), with a median of 6 days. It was smaller than 0 (mean: −2.63 days) for 27 (3.92%) pairs, meaning that the infectee showed symptoms earlier than the infector. Assuming a normal distribution (Figure 2(C)), the mean was 6.70 days (95% CI: 6.31, 7.10) and standard deviation 5.20 days (95% CI: 4.91, 5.46).

The time point of exposure was estimated from 233 pairs of infector-infectee and ranged from −9 to 13 days (Table 2), with a median of 0 day. It was smaller than 0 (mean: −3.11 days) for 102 (43.78%) pairs, meaning that the transmission occurred before the infectors showed symptoms. Assuming a normal distribution (Figure 2(D)), the mean was −0.19 days (95% CI: −0.62, 0.25) and standard deviation 3.32 days (95% CI: 2.97, 3.68).

Based on an exponential growth rate of 0.10 per day^1^ and the mean (6.7 days) and standard deviation (5.2 days) of the serial interval, R_0_ was estimated at 1.71 (95% CI: 1.67, 1.75) according to formula (1). Based on the same exponential growth rate, serial interval and mean of upper limit of latent period (2.52 days), R_0_ was estimated at 1.78 according to formula (2). Please note that the 95% CI was not available here as the 95% CI of latent period was not estimated due to limited sample size, i.e., 11 pairs of samples.

### Sensitivity and stratified and analyses

In sensitivity analysis, the estimates of incubation period, latent period and time point of exposure remained stable, with a difference of 0.14-0.3 day from those in primary analysis (Table 2), which represents the largest possible error caused by inclusion of cases with an exposure period spanning 2 or 3 days. For serial interval and time point of exposure, the results from shifted lognormal, Weibull and gamma distributions are almost the same and all of them are similar to those from normal distribution, hence only the lognormal distribution is presented (Figure 2(C)-(D) and Table 2). R_0_ remained almost the same in sensitivity analysis (data not shown).

The results of stratified analysis, where asymptomatic cases were also included, are summarized in Table 3. The three parameters were consistently associated with disease severity and generation in clusters. Specifically, for those with less severe disease or at lower generations in clusters, the incubation period, interval between their exposure and primary cases’ symptom onset, and interval between their symptom onset and primary cases’ symptom onset all tend to be longer. For those whose primary cases had less severe disease, the incubation period tends to be longer, while the interval between their symptom onset and primary cases’ symptom onset tends to be shorter. No such trend was observed for other factors.

**Table 3.** Stratified analyses of incubation period, serial interval, and time point of exposure according to selected characteristics

| Groups | Incubation period | Serial interval | Time point of exposure |
| --- | --- | --- | --- |
| Age (year) |  |  |  |
| 0 - 18 | 8.47 (6.06, 10.63) | -- | -- |
| 19 - 64 | 7.34 (6.96, 7.71) | -- | -- |
| ≥ 65 | 9.47 (8.33, 10.71) | -- | -- |
| Sex |  |  |  |
| Male | 7.24 (6.73, 7.69) | -- | -- |
| Female | 7.91 (7.43, 8.41) | -- | -- |
| Travel history to Hubei |  |  |  |
| Yes | 7.37 (6.61, 8.25) | -- | -- |
| No | 7.44 (7.03, 7.83) | -- | -- |
| Infectior was a known confirmed case origin |  |  |  |
| Yes | 7.72 (7.23, 8.21) | -- | -- |
| No | 7.10 (6.61, 7.58) | -- | -- |
| Span of exposure period |  |  |  |
| 1 day | 7.76 (7.32, 8.15) | -- | -0.02 (-0.52, 0.48) |
| 2-3 days | 6.73 (6.10, 7.28) | -- | -0.72 (-1.51, 0.11) |
| Generation of clustering cases |  |  |  |
| 1<sup>st</sup> | 5.79 (4.99, 6.63) | -- | -- |
| 2<sup>nd</sup> | 8.06 (7.55, 8.60) | 7.09 (6.62, 7.53) | 0.21 (-0.28, 0.69) |
| 3<sup>rd</sup> or higher | 6.27 (5.16, 7.33) | 5.17 (4.44, 5.96) | -1.60 (-2.33, -0.85) |
| Severity |  |  |  |
| Asymptomatic* | 11.66 (9.90, 13.30) | 9.77 (8.43, 11.21) | 1.38 (-0.13, 2.75) |
| Mild to moderate | 8.25 (7.51, 8.99) | 7.44 (6.73, 8.19) | 1.28 (0.59, 1.92) |
| Severe | 6.50 (3.86, 8.18) | 2.75 (0.19, 5.31) | -8.00 (-17.00, 1.00) |
| Severity of infectior |  |  |  |
| Asymptomatic* | 7.72 (5.33, 9.50) | -4.30 (-7.15, -1.85) | -- |
| Mild to moderate | 5.41 (4.15, 6.61) | 5.63 (4.93, 6.40) | -- |
| Severe | -- | 7.19 (5.78, 8.44) | -- |

\*In these analyses the date of diagnosis was used the day of onset for asymptomatic cases. Asymptomatic infectees were excluded from the overall analysis of incubation period and serial interval, while asymptomatic infectors were excluded from the overall analysis of serial interval and time point of exposure. --, not applicable, or not available because of too few cases.

## Discussion

By pooling individual data from seven countries, we estimated the mean incubation period of COVID-19 to be 7.44 days, latent period 2.52 days, serial interval 6.70 days, time point of exposure −0.19 day and R_0_ 1.70 to 1.78. The time point of exposure can also be inferred by mean serial interval minus mean incubation time (−0.74 day), which is consistent with the direct estimate (−0.19 day) as both suggest the mean time point of exposure to be around the day before primary cases’ symptom onset. Based on the above estimates, the timeline of infection for an ‘average’ pair of infector-infectee in a transmission chain is demonstrated in Figure 3.

**Figure 3.**
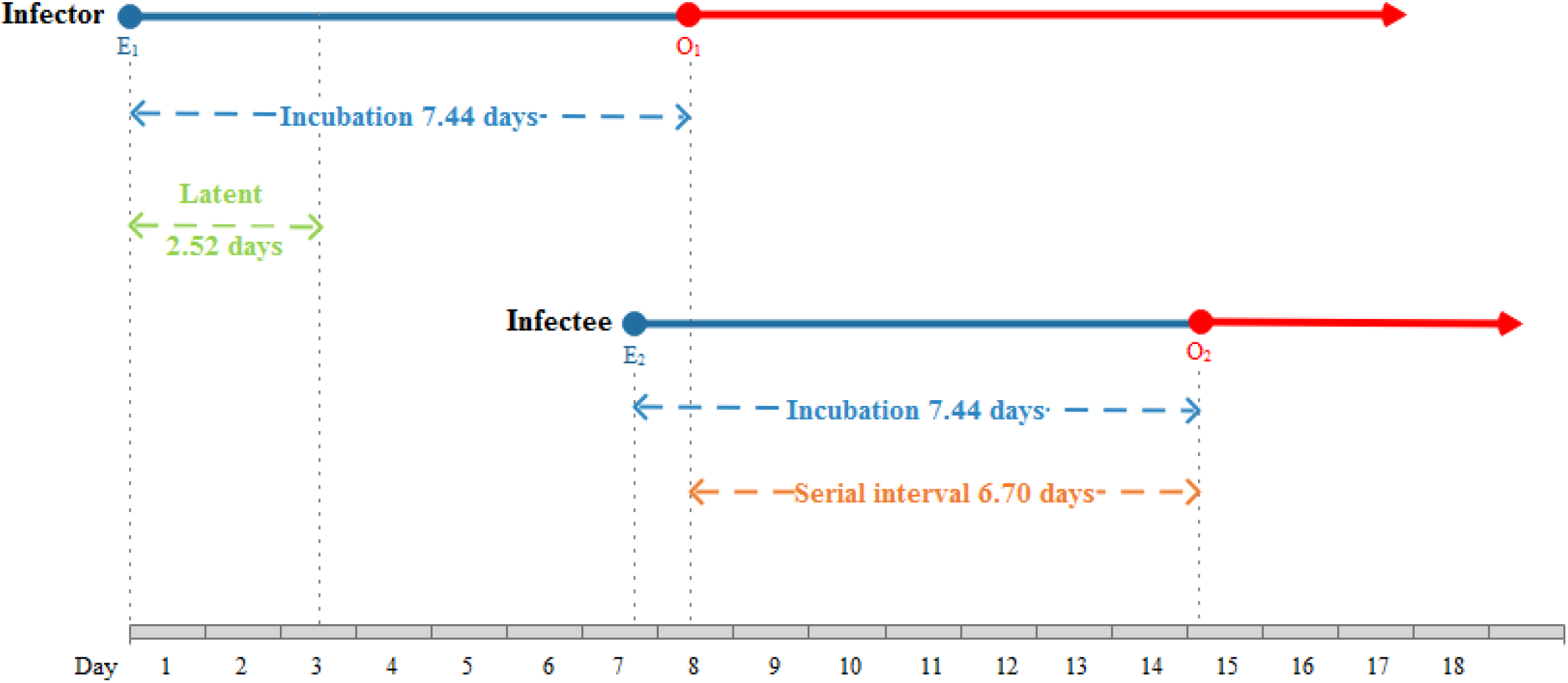
The timeline of events for an ‘average’ pair of infector-infectee in a transmission chain, according to the estimates from this study. E_1_, exposure of infector; O_1_, symptom onset of infector; E_2_, exposure of infector; O_2_, symptom onset of infector.

Our estimates of incubation period and serial interval are longer than most of the previous estimates.^2-7^ There are several possible reasons for the difference. First, the sample size is generally small in previous studies,^2,4,6,7,9-12^ but much larger in the present one. Second, most cases in previous studies had a long or even unclear interval of exposure, making it difficult to determine the exact exposure date^3,5,13^ and giving rise to error. By contrast, this study followed strict inclusion criteria regarding exposure period and method to determine exposure date to ensure the potential error in the estimates be small (< 0.3 day, according to the sensitivity analysis). Third, the approaches to determination of transmission order within clusters are different. For the cases involving cluster infection, which represent the majority of all cases, a common practice of previous studies was to take the case with earliest date of symptom onset as infector and others as infectee.^9^ However, we deemed it inappropriate in view of the varying incubation periods of individuals and potential pre-symptomatic or asymptomatic transmission. In this study, more rigorous criteria were applied to the inclusion of clusters, and the clusters in which transmission order was uncertain were excluded from relevant analyses. Fourth, different studies used data from different stages of the epidemic. The data of this study was retrieved through late February of 2020, which may reflect more of the situation after strong measures were taken to battle the epidemic in China.

Our finding that the incubation period was within 14 days for 93% of the cases lends support to current practice of 14-day quarantine of persons with potential exposure to SARS-CoV-2. In line with other studies,^13^ we also found some cases who developed symptoms 14 days after exposure, indicating that longer quarantine periods might be justified for some people. However, as it is hard to know beforehand who will develop symptoms beyond 14 days of exposure, the cost of extending quarantine of many people and the potential consequence of failure to identify a few symptomatic cases must be weighed carefully.

The negative values of serial interval and time point of exposure provide evidence of pre-symptomatic transmission, which are consistent with the much shorter latent period than incubation period. The percentages of negative serial interval (3.92%) and negative time point of exposure (43.78%) can be viewed as the lower and upper limits of probability of pre-symptomatic transmission, respectively. Asymptomatic transmission was also observed in this study (Table 3). These findings are in line with those of previous studies.^8,9,14-17^ Stratified analysis in this study found a trend towards longer incubation period of less severe cases as compared with that of more severe cases. This implies that severe cases could be detected and ‘removed’ from the pool of transmission sources more quickly, while less severe cases would stay in the pool for a longer time and constitute a big challenge to the control of epidemic given the possibility of pre-symptomatic transmission.

The R_0_ we estimated is smaller than those from previous estimates which were mostly between 2 and 4.^7,20,26-28^ As discussed above, the difference may either be due to methodological issues in obtaining parameters or reflect different stages of epidemic. In any case, a smaller R_0_ should not be interpreted as low risk of transmission. Slow response of government, pre-symptomatic and asymptomatic transmission and insufficient protection measures taken by the public together could lead to an out-of-control epidemic, as with the current situation in many countries.

This study has limitations. First, the cases were retrieved from publicly available reports, and many cases were excluded because there were no details available for estimating the parameters. Thus, the representativeness of cases could be a concern. A comparison of the age and sex of included cases with those of 72314 cases in China showed that they are generally similar.^29^ Previous studies suggested that publicly reported cases may overrepresent the severe ones. However, based on 329 cases (28.48%) with relevant information available, non-severe cases appeared to account for a higher proportion in the present study than in the previous one on 72314 cases in China. The second limitation of this study is that the upper limit of latent period was based on only 11 pairs of infector-infectee. Larger studies are needed to obtain a more robust estimate. The third limitation is that by using the upper limit of latent period instead of the real latent period in formula (2), in our case, could lead to a slightly smaller value of R_0_. If we use a naively small value of the latent period at 0 days (i.e., no latency), the formula (2) would lead to a R_0_ at 1.67, which can be treated as the lower bound. Hence, under the formula (2), R_0_ may range from 1.67 to 1.78, which was consistent with those from formula (1), i.e., 1.71 (95% CI: 1.67, 1.75).

In conclusion, this study obtained robust estimates of several key epidemiological parameters of COVID-19. It provides additional evidence on the mean incubation period of COVID-19, which supports current practice of 14-day quarantine of persons with potential exposure but also suggests that longer monitoring periods might be needed for selected groups. The estimates of serial interval, time point of exposure and latent period provide consistent evidence on pre-symptomatic transmission. Asymptomatic transmission was also observed. This together with asymptomatic transmission and the generally longer incubation and serial interval of less severe cases suggest a high risk of long-term epidemic in the absence of appropriate control measures.

## Data Availability

The data that support the findings of this study are available from the corresponding author upon reasonable request via email. After publication of study findings, the data will be available for others to request.

## Contributors

YZ conceived the idea and designed the study, has full access to all data in the study, and takes responsibility for the integrity of data and the accuracy of data analysis. MS, ZJ, ZM, YQ, GW, ZY and YZ contributed to data acquisition, statistical analysis, and writing of the report. WMH and ZS contributed to statistical analysis. All authors contributed to data interpretation and critical revision, and reviewed and approved the final version.

## Acknowledgement

We thank all the people who worked hard, risked their health and even sacrificed their lives to fight against COVID-19. They are real heroes. We also thank the numerous individuals who reduced their social activities to minimum to help contain the spread of SARS-CoV-2. Spring will surely come.

